# Motiro: an unified non-supervised framework for statistical analysis of probe-based confocal laser endomicroscopy videos of colorectal mucosa

**DOI:** 10.1101/2020.10.13.20209254

**Authors:** Alan U. Sabino, Adriana V. Safatle-Ribeiro, Fauze Maluf-Filho, Alexandre F. Ramos

**Author notes:** **Corresponding author:** Alexandre Ferreira Ramos PhD, School of Arts, Sciences and Humanities, University of São Paulo, Av. Arlindo Béttio, 1000, Ermelino Matarazzo - São Paulo, SP, 03828-000, Brazil. **Guarantor of the article:** Alexandre Ferreira Ramos. **Specific author contributions:** Videos acquisition: AVSR, FMF; Software planning and revision: AUS, AFR; Coding and software application: AUS, AFR; Results analysis: AUS, AFR; Manuscript writing: AUS, AVSR, AFR. AUS, AVSR, FMF, AFR approve the final draft submitted. **Financial support:** The study design, collection, analysis, and interpretation of the data and the manuscript was independent of the financial support. AUS thanks Coordenação de Aperfeiçoamento de Pessoal de Nível Superior - Brasil (CAPES) Finance Code 88882.376562/2019-01 for partial financial support of this study. AFR thanks CAPES Finance Code 88881.062174/2014-01 for partial support of this study. **Potential competing interests:** The authors declare no potential competing interest.

## Abstract

**Objective:** To present Motiro, an unified framework for non-supervised statistical analysis endomicroscopy videos of the colorectal mucosa.

**Materials and Methods:** We wrote an open-source Python wrapper using ImageJ software with OpenCV, Seaborn and NumPy libraries. It generates a mosaic from the video of the mucosa, evaluates morphometric properties of the crypts, their distribution, and return their statistics. Shannon entropy (and Hellinger distance) are used for quantifying variability (and comparing different mucosa).

**Results:** The segmentation process applied to normal mucosa of pre(post)- neoadjuvant patient is presented along with the corresponding statistical analysis of morphometric parameters.

**Discussion:** Our analysis provides estimation of morphometric parameters consistent with available methods, is faster, and, additionally, provides statistical characterization of the mucosa morphometry. Motiro enables the analysis of large amounts of endomicroscopy videos for building a normal rectum features dataset to help on: detection of small variability; classification of post-neoadjuvant recovery; decision about surgical intervention necessity.

## INTRODUCTION

Probe-based laser endomicroscopy (pCLE) of colorectal cancer (CRC) enhanced screening and post-neoadjuvant surveillance(1). pCLE videos aid endoscopists to classify population groups accordingly with their probabilities of developing a CRC(2), *e.g*. by aberrant crypt *foci* quantification for inference of recurrence(3) and neoplasms(4) chances. That entails the use of computational tools for complementing human analysis(5) using machine learning-based classification of polyps(6) or quantitative analysis of the mucosa architecture by supervised, decoupled use of Icy and ImageJ softwares(7). In Ref.(7), the necessity of a unified framework for the analysis of the architecture of the mucosa was raised. In this manuscript we present Motiro^1^, a Python-based non-supervised unified framework for statistically characterize the morphometry of the colorectal mucosa at pre or post neoadjuvant CRC using pCLE videos. Because of its unified, non-supervised functioning, Motiro enables the efficient creation of a database of analyzed images and clinical data(8) to be jointly used for elaborating an optimal screening and surveillance agenda for CRC patients(2,9,10).

The molecular processes controlling tissue patterning in the colorectal mucosa(11) are unavoidable noisy(12). That may cause small fluctuations on the position, shape, and size of the crypts of the normal mucosa. Indeed, the link between molecular level fluctuations and tissue level variation has been reported on analysis of *Drosophila* embryos development(13,14) and it is fair to extrapolate such a conceptual framework for understanding the patterning of the colorectal mucosa. Therefore, we analyze the morphometric parameters of the crypts using histograms to represent the small degree of disorder exhibited by the normal mucosa. For distinguishing the disorder of normal mucosa from that observed at earlier stages of a neoplasm or recurrent CRC, Motiro brings two methods for evaluating the distribution of morphometric parameters, namely, the differential Shannon entropy and the Hellinger distance. The former has been widely used to quantify the disorder in physical systems(15), and the latter is employed to compare the overlay of two histograms (16) representing the distribution of a given morphometric parameter. These two quantities enable one to comparatively compute the degree of disorder of the mucosa of, for example, a patient and a population, the crypts at pre- and post-neoadjuvant therapy, or different regions of the mucosa of a patient. They also set a quantitative criterion for helping on the distinction of normal-abnormal variability of the mucosa’s architecture. Moreover, the unsupervised functioning of Motiro enables one to efficiently construct a large database of endomicroscopic images statistically characterized. That has a clear clinical implication: one may quantify the probability of an observed variability within the colon mucosa of a patient to be the classified as either normal or as the early stage of a neoplasm or recurrent CRC by, *e.g*., application of a hypothesis test.

## METHODS

### Dataset images

The images were acquired using pCLE. pCLE is a real time *in vivo* method for acquisition of 1000 times magnified optical biopsies for evaluating cellular and vascular patterns. Before the pCLE procedure, 5 ml of 10% fluorescein diluted in 100 ml of saline solution were injected intravenously. The probe was inserted through the working channel of the endoscope into the rectum a few minutes after the fluorescein injection. The pCLE (2.5 mm UHD ColoFlex probe, Cellvizio; Mauna Kea Technologies, Paris, France) provided depth of examination of 55 to 65 μm, a 240 μm field of view at a resolution of 1 μm and magnification of 1000X at 12 frames/s. In all patients, normal mucosa located at least 5 cm from the target lesion was also examined by pCLE, in order to have a comparison image to the altered mucosa.

### Framework software

Figure 1 shows a flowchart of the major stages of Motiro. We use a pCLE video as input of our wrapper(17). Stage 1, Motiro combines tools from Open Source Computer Vision Library (OpenCV)(18), and ImageJ plugin Register Virtual Stack Slices (RVSS)(19,20). OpenCV (RVSS) is used for dismantling the video into frames and text removal (frame stitching). On the resulting mosaic, OpenCV tools are employed for *pre-processing, segmentation using k-means algorithm(21)*, and *morphometric* analysis are executed in Stage 2 (please, see Supplementary Digital Content 1 (SDC1) for a detailed description). Stage 3, NumPy and Seaborn Python libraries are used for *statistical analysis*, and data generation for calculation of differential Shannon entropy and Hellinger distance.

**Figure 1:**
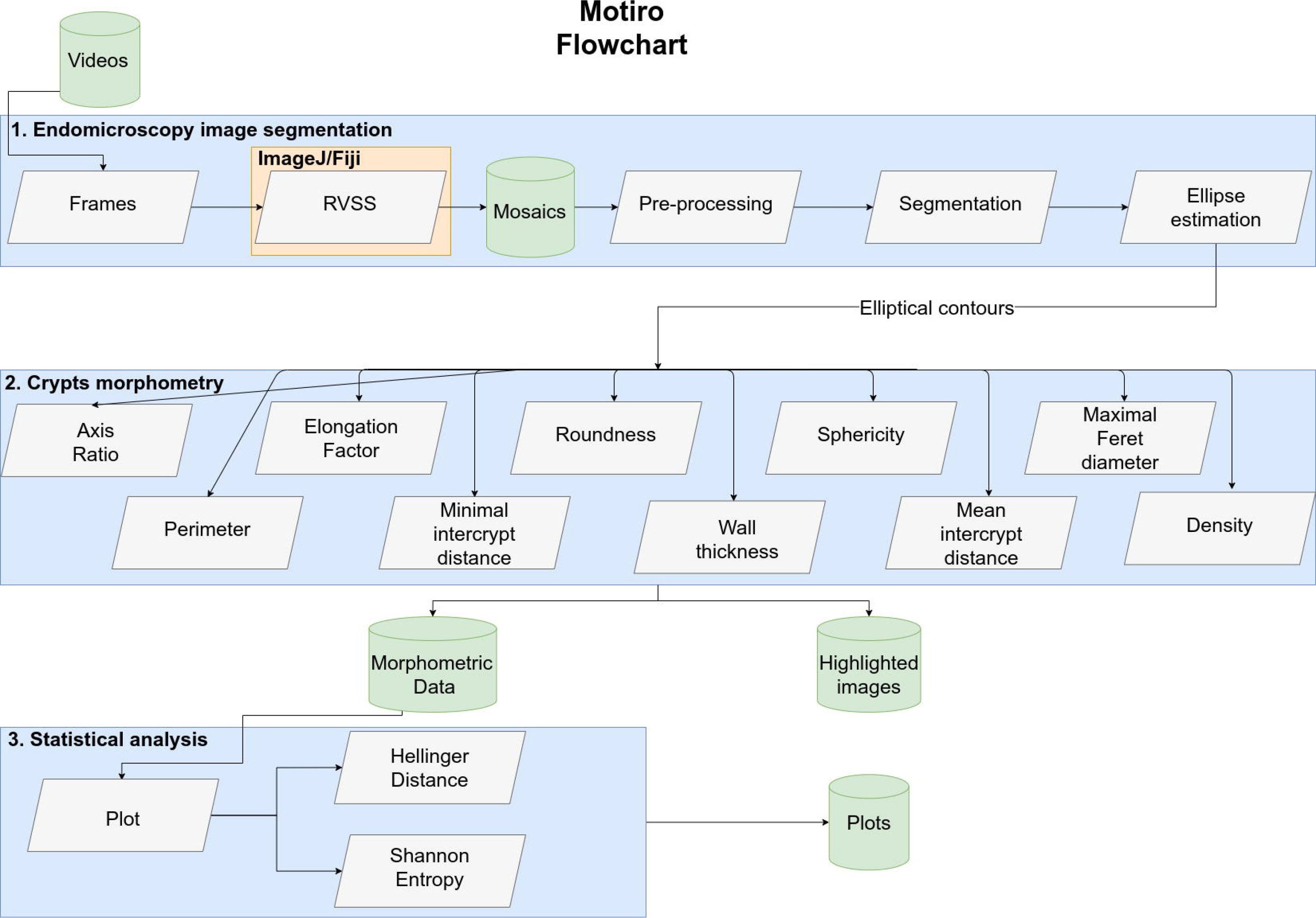
Motiro flowchart. A visual representation of the major stages of Motiro. Stage 1 receives endomicroscopic videos as input and provides mosaics and the elliptical contours as results. Stage 2 the contours are used to analyze the morphometric data and highlight the images for visual assessment of the parameters estimation. Stage 3 generates the output by plotting the statistical analysis of the morphometric parameters.

### Morphometric parameters

A geometric interpretation of our results was facilitated by converting pixel values on the mosaic to micrometers. The crypts were approximated as elliptical contours (here on denoted simply as contours) drawn using OpenCV. The ratio of the major to the minor edges of a rectangle parallel to the Cartesian axis surrounding the elliptical crypt is the axis ratio (*α*). The elongation factor (*ε*), is that ratio in a rotated rectangle with edges parallel to the major (2 *a*) and minor (2 *b*) axis of the contour, with the former, called maximal Feret diameter, being estimated by both an *exhaustive search* and a *heuristic* algorithm. The perimeter (*ρ*) of the contour is estimated using(22):

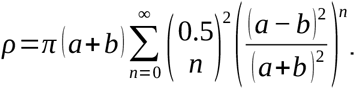

The area of a contour (*A*) is estimated in μm^2^ using, *A*=*π a b* with roundness (*Σ*) being:

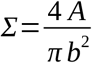

and sphericity (*σ*) being:

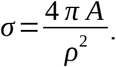

The mucosa state is further characterized by the crypts’ distribution. We estimate the mean (*Δ*) and minimal (*δ*) intercrypt distances; the minimal distance separating two nearest neighbor contours, called wall thickness (*ω*), and the tissue density (*θ*). Please, see SDC2 for further details on the methods of analysis of the morphometric parameters.

### Statistical analysis

The images were classified as pre (R) and post (T) neoadjuvant for a statistical evaluation of the morphometric parameters. We quantify the degree of disorganization of R and T images applying the differential Shannon entropy(23) on the histogram of each morphometric parameter:

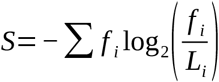

where *f_i_* is the relative frequency of observing the *i*-th range of values of a morphometric parameter and *L*_*i*_ is the *i*-th bin width (please, see SDC3 for further details). The differential Shannon entropy for a uniform distribution of a continuous random variable within an interval of width *L* is *S*_0_=log_2_ (*L*) can be used as a reference value determining the maximal degree of disorder of a distribution or histogram. Then we define the quantity 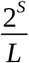 to evaluate the disorder of a distribution or histogram relative to its uniform counterpart: the distribution or histogram reflects a higher degree of disorder for 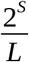 and better organization otherwise.

The use of histograms for characterizing the morphometric parameters requires the use of a statistical distance for differentiating two sets of images. Here we choose the Hellinger distance(16,24,25) between two distributions *P* and *Q*, denoted by *H* (*P, Q*), to compute the overlay between two probability densities:

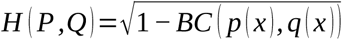

where *p* (*x*) and *q* (*x*) are probability densities evaluated in *x* and

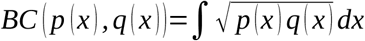

is the Bhattacharyya coefficient. Relative frequencies are used to approximate the probability densities when we compare the histograms of a morphometric parameter from two images, R and T (please, see SDC3 for further details).

## RESULTS

### Endomicroscopy image segmentation

*Figure 2* shows an example of the Motiro estimation of the crypts’ contours as obtained from the pCLE videos. *Fig 2A* shows a mosaic which irregular geometry results from the imaging acquisition process: the sensibility of the probe and absence of reference points causes a maneuvering variability. The field of view of the probe is wider than the diameter of a crypt in a normal mucosa which leads to a mosaic composed by multiple crypts prone to statistical analysis. Application of contrast enhancement and noise removal highlights the crypts from background as shown in *Fig. 2B. Fig 2C* shows a segmentation of the crypts and surrounding stroma. Application of morphological operations and application of convex hull to smooth the crypts’ peripheries is shown in *Fig 2D. Fig. 2E* shows the elliptical contours estimation of the crypts’ surrounding after the convex hull. Inspection of Figs 2E and 2C indicates the similarity of the elliptical estimates to the segmented crypts after removal of noise of the stroma and irregularities of the crypts’ boundaries. *Fig. 2F* shows Motiro’s (Icy’s(26)) contours estimation in green (red) overlaid to the original mosaic. The contours of some crypts are less accurate because of inaccuracy of segmentation process caused by the brightness of some crypts border being similar to that of the background.

**Figure 2:**
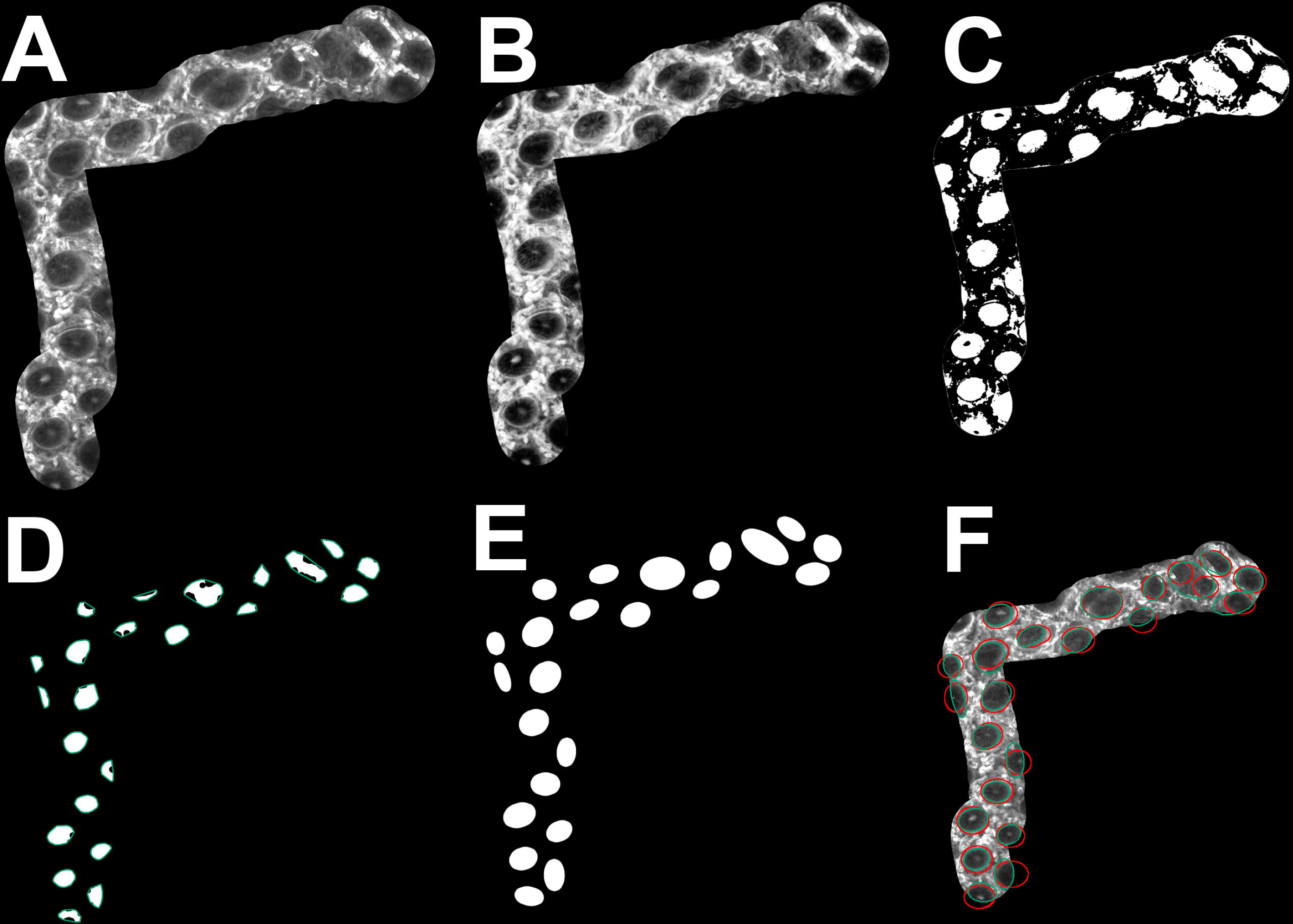
Crypts estimation process. (A) Mosaic image built from by pCLE video. (B) Mosaic after noise removal and enhanced contrast processes. (C) Binary image after application of threshold of pixels clustered in the darkest groups. (D) Result of morphological and convex hull (green boundary) operations applied on segmented image. (E) Estimated ellipses after application of convex hull. (F) Overlay of rough mosaic image, estimated elliptic contour obtained using Motiro (green), and Icy-based algorithm(red).

### Statistical analysis of the crypts morphometry

*Fig. 3* shows our statistical analysis, the superposed blue and beige histograms summarize data for R and T images, respectively, after outliers’ removal (please, see SDC4 for complete data analysis). The prevalence of single mode histograms and similarity between R and T images indicate the regular structure of the normal mucosa. Most of the axis ratio (and elongation factor) of the crypts lie within the range [1, 1.37] (and [1,1.4]) embracing 84% (and 78%) of the crypts in the R and 75% (and 69%) in the T images. The R and T images concentrate 74% and 78%, respectively, of the crypts’ roundness (and 84% and 81% for sphericity) within the range of [66%,90%] (and [94.6%,99.9%]). The maximal Feret diameter (and the perimeter) of the crypts are lying within the ranges of [81,146] (and [229,461]) embracing 98% (and 98%) in the R and 92% (and 89%) in the T images. The minimal intercrypt distance (and the wall thickness) are lying within the range [104,155] (and [4,48]) that encompass 84% (and 71%) in the R images and 81% (and 93%) in the T ones. The mean intercrypt distance values are distributed accordingly with histograms having a mode within the range [118, 180] and another within [211, 273] which, respectively, in the R (and T) images concentrate 56% (and 33%) and another 30,3% (37,5%) of the data.

**Figure 3:**
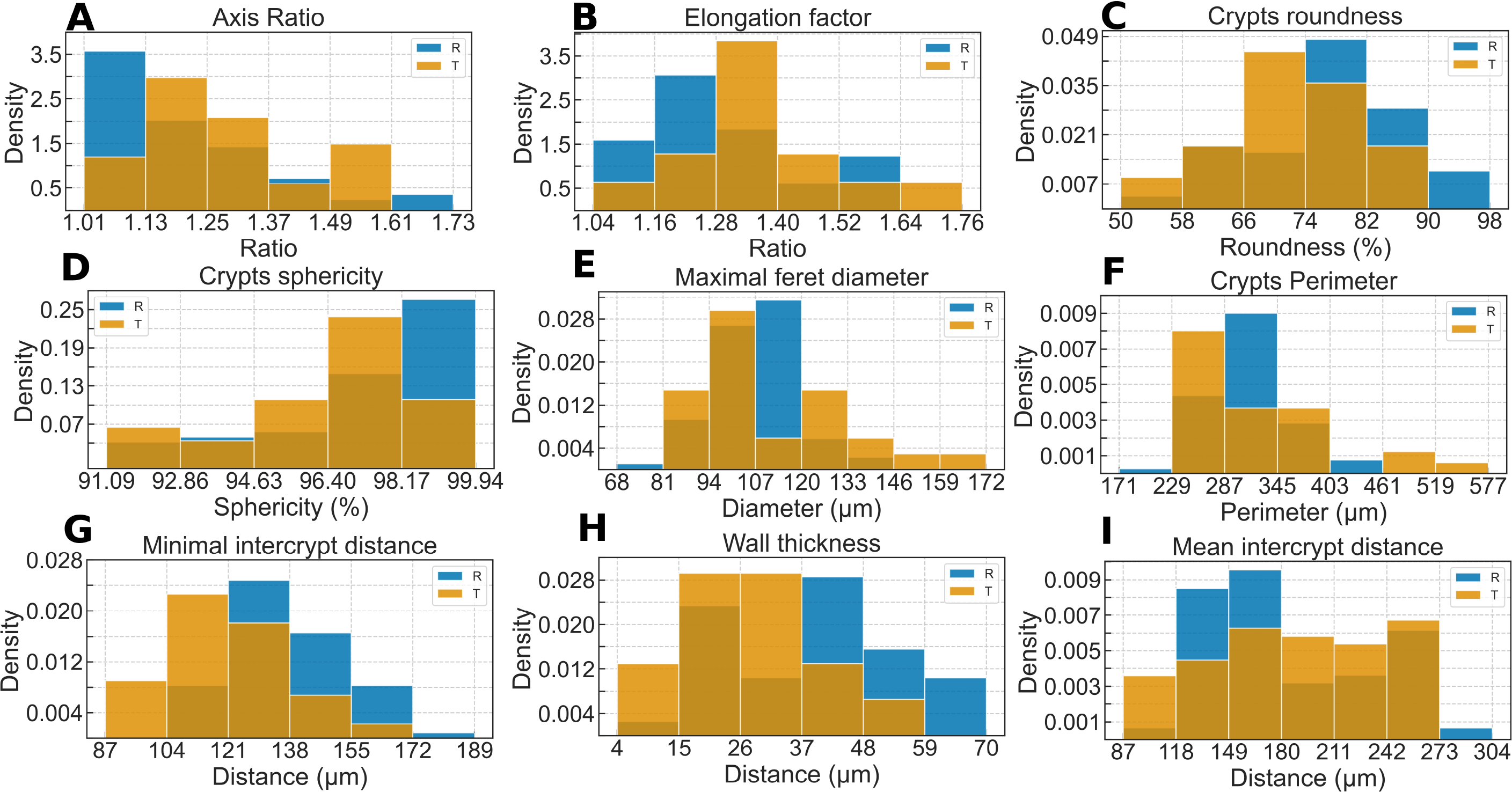
Morphometry parameters evaluation. The blue and beige histograms represent the statistics of the mucosa’s morphometric parameters in R and T neoadjuvant treatment, respectively.

Table 1 gives additional statistical information, after outliers’ removal, with columns 3 to 9 labeled as morphometric parameters as in Figs. 3A to 3I. The first two rows account for analysis of images R and T, with their first sub-row indicating mean values and standard deviations and the degree of disorder given in the second sub-row. The third row shows the Hellinger distance between histograms from R and T images. The Hellinger distance ranges from 0.230 to 0.407 and it is fair to conclude that there is a good similarity on the morphometry of the normal mucosa in both R and T images. Such a conclusion is reinforced by noticing that, for almost all morphometric parameters, the absolute value of the difference between the means of R and T images lay within the standard deviation of one or another, the exception being *δ*, which standard deviation in the R images is smaller. We compare the differential entropy of each histogram to their corresponding uniform and 2^*S*^ / *L*< 0.848. That reinforces our conclusion that the evaluated mucosa has a high degree of order despite their intrinsic variability.

**TABLE 1:** Statistical analysis of R and T images.

| | | Axis Ratio | Elongation Factor | Roundness(%) | Sphericity(%) | Maximal Feret diameter($\mu\text{m}$) | Perimeter($\mu\text{m}$) | Minimal intercrypt distance($\mu\text{m}$) | Wall thickness($\mu\text{m}$) | Mean intercrypt distance($\mu\text{m}$) |
| --- | --- | --- | --- | --- | --- | --- | --- | --- | --- | --- |
| R | mean ($\pm\text{std}$) | 1.20 ($\pm 0.16$) | 1.29 ($\pm 0.1$) | 77.4 ($\pm 9.8$) | 97.1 ($\pm 2.3$) | 108.3 ($\pm 12.3$) | 310.2 ($\pm 42.6$) | 137.9 ($\pm 15.1$) | 39.4 ($\pm 15.7$) | 185.2 ($\pm 47.5$) |
|  | Degree of disorder | 0.715 | 0.741 | 0.792 | 0.769 | 0.484 | 0.468 | 0.610 | 0.836 | 0.748 |
| T | mean ($\pm\text{std}$) | 1.28 ($\pm 0.14$) | 1.36 ($\pm 0.13$) | 72.2 ($\pm 9.1$) | 96.2 ($\pm 2.2$) | 111.0 ($\pm 19.7$) | 320.9 ($\pm 74.5$) | 120.5 ($\pm 18.8$) | 26.8 ($\pm 13.1$) | 189.6 ($\pm 52.1$) |
|  | Degree of disorder | 0.738 | 0.766 | 0.725 | 0.848 | 0.649 | 0.537 | 0.671 | 0.728 | 0.840 |
|  | Statistical Distance | 0.318 | 0.330 | 0.292 | 0.222 | 0.367 | 0.367 | 0.407 | 0.367 | 0.230 |

## DISCUSSION

The morphometry analysis presented here has been performed by Quénéhervé(7) and collaborators(QA) using Icy and ImageJ softwares separately. Because in QA approach the crypts are contoured manually, one may consider these contours as the gold-standard. In SDC5 we present a quantitative comparison between Motiro and QA morphometric estimates and obtain a mean relative error of 0.167. That demonstrates the viability of the unified non-supervised segmentation of pCLE videos of the colon’s mucosa.

Though the learning curves of Motiro and QA are similar, Motiro unifies functionalities, adds new functionalities, is non-supervised and is 5.7 times faster than QA with heuristic algorithm to evaluate crypts(SD6). Motiro runs on a Linux Operational System, requires installation of Python(3.6.9), Seaborn(0.10.1), Numpy(1.19), and OpenCV(4.3.0), and is executed in a terminal (instead of a graphical interface). Images with aberrant crypts could not be properly segmented by Motiro and in future work we expect to work on those drawbacks.

The addition of the statistical analysis of the morphometric parameters enables the quantification of the intrinsic small fluctuations of the mucosa’s architecture. For such study, we interpret the crypt-crypt interlinks as edges of a graph, which enables us to establish an estimate of the topographical properties of the mucosa. Then, we can use the differential Shannon entropy for quantifying the degree of disorder of the mucosa, and Hellinger distances for comparing the statistics of the architecture of the multiple mucosas, or of different positions or instants of the same mucosa. The statistical analysis based on removed outliers helps to reduce the bias caused by the analysis of partial crypts appearing within the edges of the mosaics. The use of histogram-based statistical analysis (as alternative to often employed average, mode, and median) opens the way for a comprehensive approach based on machine learning techniques for the classification of the normal mucosa in R and T images and estimation of neoadjuvance success chances. The analysis of a large data set may indicate whether the histogram-level differences on morphometric parameters of R and T images have a significance and set reference values for both the differential Shannon entropy and the Hellinger distances.

Motiro brings significant improvements to statistically assess and analyze colon morphometry using pCLE videos, and is ready for approaching a large amount of data obtained from normal and quasi-normal mucosa. Motiro’s image analysis approach has the benefit of not needing a large amount of pre-classified data to extract features and the morphometry parameters have clear geometrical and biological interpretations. That contrasts with machine learning approaches which demand large amounts of high-quality data to provide models which geometrical or biological meaning can be hard to determine(27). Therefore, machine learning approaches may benefit from the construction of a set of results having a clear interpretation. Indeed, Motiro is prompt to be employed for building a large database of mucosa features to be used on artificial intelligence studies aiming to establish the connection among mucosa morphometry and clinical data. The use of statistical analysis enables a refinement on the differentiation of normal and post-neoadjuvant fully recovered mucosa. That may help on elaborating more assertive machine learning-based models to assist physicians to decide about the necessity of surgical interventions on post-neoadjuvant colorectal cancer patients. Additionally, the use of a unified framework for a quantitative characterization of the architecture of the mucosa enables to set additional standards to aid human analysis by reducing the role of subjectivity(28). Besides, the modular structure of Motiro can be used for its adaptation for analyzing images obtained by advanced methods(29).

## Supporting information

Supplementary Digital Content

## Data Availability

The authors confirm that the data supporting the findings of this study are available within the article and its supplementary materials.

## ACKNOWLEDGMENTS

We would like to thank Prof. Roger Chammas for helpful comments.

## SUPPLEMENTARY MATERIAL

Supplementary material is available online.

## Footnotes

1 From tupi-guarani, the language of native Brazilians, meaning *a reunion for building*.

## REFERENCES

1. Kiesslich R, Burg J, Vieth M, et al. Confocal laser endoscopy for diagnosing intraepithelial neoplasias and colorectal cancer in vivo. Gastroenterology 2004;127:706–713.

2. Gupta S, Lieberman D, Anderson JC, et al. Recommendations for Follow-Up After Colonoscopy and Polypectomy: A Consensus Update by the US Multi-Society Task Force on Colorectal Cancer. Gastroenterology 2020;158:1131-1153.e5.

3. Uchiyama T, Takahashi H, Endo H, et al. Number of aberrant crypt foci in the rectum is a useful surrogate marker of colorectal adenoma recurrence. Dig. Endosc. Off. J. Jpn. Gastroenterol. Endosc. Soc. 2012;24:353–357.

4. Kowalczyk M, Orłowski M, Klepacki Ł, et al. Rectal aberrant crypt foci (ACF) as a predictor of benign and malignant neoplastic lesions in the large intestine. BMC Cancer 2020;20:133.

5. Karstensen JG. Evaluation of confocal laser endomicroscopy for assessment and monitoring of therapeutic response in patients with inflammatory bowel disease. Dan. Med. J. 2016;63

6. André B, Vercauteren T, Buchner AM, et al. Software for automated classification of probe-based confocal laser endomicroscopy videos of colorectal polyps. World J. Gastroenterol. 2012;18:5560–5569.

7. Quénéhervé L, David G, Bourreille A, et al. Quantitative assessment of mucosal architecture using computer-based analysis of confocal laser endomicroscopy in inflammatory bowel diseases. Gastrointest. Endosc. 2018;89:626–636.

8. Kowalczyk M, Orłowski M, Siermontowski P, et al. Occurrence of colorectal aberrant crypt foci depending on age and dietary patterns of patients. BMC Cancer 2018;18:213.

9. Dey N, Kochman ML, Komanduri S, et al. Report from the AGA Center for GI Innovation and Technology’s Consensus Conference: Envisioning Next-Generation Paradigms in Colorectal Cancer Screening and Surveillance. Gastroenterology 2020;158:455–460.

10. Melson JE, Imperiale TF, Itzkowitz SH, et al. AGA White Paper: Roadmap for the Future of Colorectal Cancer Screening in the United States. Clin. Gastroenterol. Hepatol. 2020;S1542-3565(20)30917–4

11. Murray PJ, Kang J-W, Mirams GR, et al. Modeling Spatially Regulated β-Catenin Dynamics and Invasion in Intestinal Crypts. Biophys. J. 2010;99:716–725.

12. Delbrück M. Statistical Fluctuations in Autocatalytic Reactions. J. Chem. Phys. 1940;8:120–124.

13. Surkova S, Kosman D, Kozlov K, et al. Characterization of the Drosophila segment determination morphome. Dev. Biol. 2008;313:844–862.

14. Prata GN, Hornos JEM, Ramos AF. Stochastic model for gene transcription on Drosophila melanogaster embryos. Phys. Rev. E 2016;93:022403.

15. Faraggi E, Dunker AK, Jernigan RL, et al. Entropy, Fluctuations, and Disordered Proteins. Entropy 2019;21:764.

16. Deza MM, Deza E. Encyclopedia of Distances. Berlin, Heidelberg: Springer Berlin Heidelberg; 2009.

17. Applied Mathematics and Biological Physics Laboratory: Endomicroscopy analysis [Internet]. [cited 2020 Jul 8] Available from: https://github.com/amphybio/endomicroscopy-analysis

18. Bradski G, Kaehler A. OpenCV. Dr Dobb’s J. Softw. Tools 2000;3

19. Arganda-Carreras I, Sorzano COS, Marabini R, et al. Consistent and Elastic Registration of Histological Sections Using Vector-Spline Regularization. In: Beichel RR, Sonka M, editor(s). Computer Vision Approaches to Medical Image Analysis. Berlin, Heidelberg: Springer Berlin Heidelberg; 2006. p. 85–95.

20. Schneider CA, Rasband WS, Eliceiri KW. NIH Image to ImageJ: 25 years of image analysis. Nat. Methods 2012;9:671–675.

21. Jain AK, Murty MN, Flynn PJ. Data clustering: a review. ACM Comput. Surv. 1999;31:264–323.

22. Ivory J. VIII. A New Series for the Rectification of the Ellipsis; together with some Observations on the Evolution of the Formula (a2 + b2 - 2ab cos λ). Earth Environ. Sci. Trans. R. Soc. Edinb. 1798;4:177–190.

23. Cover TM, Thomas JA. Elements of information theory. 2nd ed. Hoboken, N.J: Wiley-Interscience; 2006.

24. Chung JK, Kannappan PL, Ng CT, et al. Measures of distance between probability distributions. J. Math. Anal. Appl. 1989;138:280–292.

25. Gibbs AL, Su FE. On Choosing and Bounding Probability Metrics. Int. Stat. Rev. 2002;70:419–435.

26. Chaumont F de, Dallongeville S, Chenouard N, et al. Icy: an open bioimage informatics platform for extended reproducible research. Nat. Methods 2012;9:690–696.

27. Yu K-H, Beam AL, Kohane IS. Artificial intelligence in healthcare. Nat. Biomed. Eng. 2018;2:719–731.

28. Chang J, Ip M, Yang M, et al. The learning curve, interobserver, and intraobserver agreement of endoscopic confocal laser endomicroscopy in the assessment of mucosal barrier defects. Gastrointest. Endosc. 2016;83:785-791.e1.

29. Prieto SP, Reed CL, James HM, et al. Differences in colonic crypt morphology of spontaneous and colitis-associated murine models via second harmonic generation imaging to quantify colon cancer development. BMC Cancer 2019;19:428.

