## Supplementary Digital Content for "Motiro: an unified non-supervised framework for statistical analysis of probe-based confocal laser endomicroscopy videos of colorectal mucosa"

Further details on implementing Motiro and its evaluation in comparison with previously available approaches is given in this supplementary digital content. Section 1 gives an overview of the framework and the steps needed for segmentation. Section 2 further discusses the morphometric parameters, comparison of the runtime of the algorithms for the data analyzed here, the precision of the algorithms used here and the distinct methods for perimeter and area estimation. Further description of the statistical analysis is given in Section 3. The histograms and statistical analysis with complete data are presented in Section 4. An analysis comparing the results and runtime benchmark obtained with Motiro to previous approach by Quénéhervé and collaborators is given in Sections 5 and 6.

1. **Framework software**

Motiro framework analyses pCLE videos of the colorectal mucosa in the following major steps(1): i) we use OpenCV(2) to dismantle the endomicroscopic videos into frames and for text information removal; ii) we use RVSS(3) to create a mosaic image by stitching the frames together. This tool employs the difference of Gaussian detector to identify feature points in the images and the scale-invariant feature transform algorithm to match common points between subsequent frames(4); iii) we use OpenCV tools to perform the morphometric analysis of the mosaic image in four stages:

*1. pre-processing and segmentation*: we use $k$-means clustering algorithm(5) to separate the crypts from the background, and, on the resulting image, we apply an ellipse shaped structuring element to execute erosion and dilatation for enhancing the crypt's morphology; The contrast image was enhanced by global histogram equalization and Gaussian blur was used for noise removal. Pixels of similar fluorescence intensities were clustered setting $k$-means for four groups, the two darkest indicating the crypts' centers and their peripheral regions, and the outer regions are given by the lighter pixels.

*2. ellipse estimation*: on each pre-processed crypt we apply the convex hull operation to remove irregularities and use the result to generate an approximating elliptical contour for the original crypts. That is useful because of finger-like irregularities formed within the borders of the crypts as a consequence of the non-uniform fluorescence intensity distribution within a crypt as it can be noticed on the mosaic in the article Fig. 2A. The application of the convex hull enables the removal of those irregularities and the smoothing of contour of the crypts. The approximation of the contours by ellipses is the final step on the preparation for the analysis of the morphometry of the mucosa.

*3. morphometrical analysis:* in each elliptical contour of the segmented images we analyze the axis ratio, elongation factor, maximal Feret diameter, perimeter, roundness, sphericity, as well as, the mean and minimal intercrypts distance, wall thickness and tissue density.

*4. statistical analysis:* we build histograms of the distribution of values of the morphometric parameters defined on step 3 on pre(R) and post(T)-neoadjuvant images of non-tumoral regions of the tumor. Then, the disorder of each morphometric parameter is estimated using the differential Shannon entropy. A comparison between the histograms for a given morphometric parameter on a R and T mosaics is done using the Hellinger distance.

2. **Morphometric parameters**

The mosaic enables the analysis of an amplified stationary image of the targeted tissue area and a quantitative evaluation of its morphological properties. Pixel values were converted to micrometers to enable a geometric interpretation of the results: 51 pixels on the image corresponds to a length of 20μm. The elliptical contours of crypts were draw using OpenCV library. A rectangle surrounding the elliptical crypt is draw with its basis being parallel to the horizontal axis, and the *ratio of its major to minor axis* is denoted by $\alpha$. We also defined a rotated rectangle enclosing the elliptical crypt and used the ratio of its major ($2a$) and minor ($2b$) axis to define the *elongation factor*, denoted by $\varepsilon$. We denote the longest line connecting two points of the elliptical crypts, called *maximal Feret diameter*, by $2a$ where $a$ is the semi-major axis of the estimated ellipse. The estimation of maximal Feret diameter was done using both an exhaustive search and a heuristic algorithm. In the exhaustive search the maximal Feret diameter is estimated as the length of the largest of the edges connecting all pairs of pixels of the elliptic contour of the crypts. The heuristic estimation takes the largest value between $x_{2}-x_{1}$ and $y_{2}-y_{1}$ where $x_{1}$ and $y_{1}$ (or $x_{2}$ and $y_{2}$) are minimal (or maximal) positions of the pixels of the contour of the crypt on the horizontal and vertical axis, respectively.

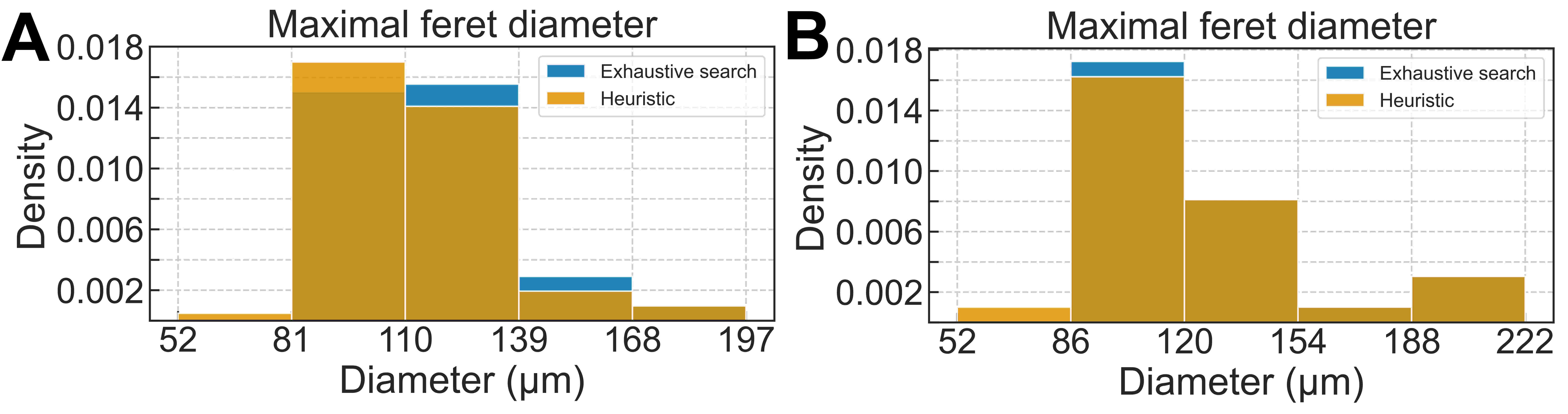
**Supplementary Figure** 1: **Maximal Feret diameter estimation.** Estimated value with exhaustive search and heuristic algorithm for R **(A)** and T **(B)** images

Supplementary Fig. 1 shows the distribution of values of the maximal Feret diameter in R and T images. The histograms are very similar with the heuristic algorithm presenting a small bias of underestimating the maximal Feret diameters. The Hellinger distance between R and T histograms in Supplementary Figs 1A and 1B are, respectively, 0.099 and 0.132. For R (and T) images the mean value ± standard deviation of the maximal Feret diameters were estimated using both methods. The heuristic method gives 112.04 μm ± 19.39 μm (and 121.37 μm ±35.64 μm) and the exhaustive search gives 115.60 μm ±19.92 μm (and 122.81 μm ±36.28 μm). We define the relative error as $\left| e-h \right|/M$, where $e$ (and $h$) is the estimated value using the exhaustive search (and heuristic) algorithm for maximal Feret diameter and $M=\left( e+h \right)/2$. The results on errors being 0.03 for R and 0.01 for T image. The time elapsed to run the heuristic function was 1 second while with exhaustive search was 202 seconds. Analyzing the mean estimated values, the Hellinger distance and the small relative error we can verify that the algorithms produce sufficiently similar estimations. The heuristic algorithm provides a two hundred times faster performance and may be very useful when we employ Motiro on the evaluation of large amounts of images.

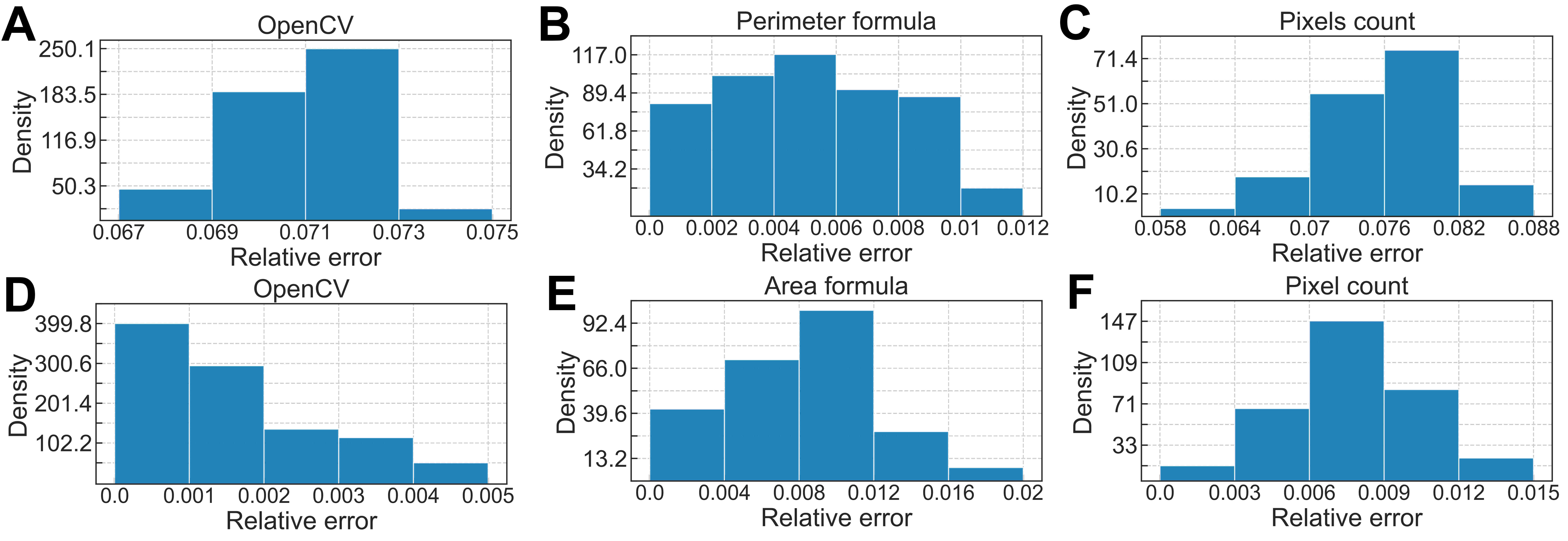

 **Supplementary Figure 2: Ellipse perimeter and area relative error distribution**. **(A-C)** Ellipse perimeter (and area **(D-F)**) relative error distribution of estimated crypts using OpenCV **(A and D)**, perimeter (area) formula **(B and E)** and pixels count **(C and F)**.

There are multiple methods for estimating the perimeter and area of an ellipse in an image, and we choose three of them: the OpenCV implementation, the formulas presented on the methods section of the main text, and pixels count. The histograms in *Supplementary Figure 2* show the relative error of the estimation of the $i$-th ellipse perimeter and area on the first and second row, respectively. In each histogram, the relative error is given by $\left| e_{i}-\mu_{M} \right|/{\mu_{M}}$, where $e_{i}$ is the estimated value for perimeter (area) of the $i$-th ellipse for each method and $\mu_{M}$ is the mean value among OpenCV, perimeter (area) formula and pixels count. The relative error of the perimeter with OpenCV (and pixels count) concentrates 96.6% (and 89.2%) of the errors observed within the range [0.067; 0.073] (and [0.064; 0.082]) while the perimeter formula concentrates 96% within the range [0; 0.01]. The relative error of the area with OpenCV (and pixels count) are lying within the range [0; 0.004] (and [0.003; 0.012]) embracing 94.7% (and 89,5%) whereas the area formula lie within the range [0; 0.012] embracing 85.3% of the errors, there is not a gap among the error distributions of methods. Assuming that the mean estimate $\mu_{M}$ is closer to the actual parameter value, it is fair to conclude that for both the perimeter and area, the formulas give a sufficiently good estimate while ensuring fast processing times.

***Supplementary Table 1:*** *Summary of perimeter and area estimation with different methods.*

|  | **OpenCV**  **mean (± std)** | **Formula**  **mean (± std)** | **Pixels count**  **mean (± std)** |
| --- | --- | --- | --- |
| **Perimeter (μm)** | 343.75 (± 71.08) | 321.27 (± 64) | 296.73 (± 60.52) |
| **Area (μm²)** | 8233.22 (± 3334.21) | 8183.11 (± 3364.49) | 8291.59 (± 3345.86) |

The *Supplementary Table 1* presents the mean and standard deviation (within parenthesis) of the perimeter and area of the ellipses as estimated using the three methods. The absolute value of the difference between the means of each method are within the standard deviation of any other. That reinforces the applicability of our choice for the formulas for estimating the perimeter and areas.

The roundness of an elliptical crypt, denoted by $\Sigma$, is:

$\Sigma=\frac{4A}{\pi b^{2}}$

and its sphericity, denoted by $\sigma$, is:

$\sigma=\frac{4\pi A}{\rho^{2}}$

and those quantities help us to determine the regularity of the crypts morphometry within a given mucosa.

Besides the crypts morphology, their distribution within the analyzed region is important to determine the condition of the mucosa. The mean and minimal intercrypt distance, denoted by $\Delta$ and $\delta$, are, respectively, given by the average and the least distance between the center of an elliptical crypt and their neighbors centers. The wall thickness, denoted by $\omega$, is the minimal distance separating the contour of two nearest neighbor elliptical contours. Either exhaustive search and heuristic algorithms were employed for estimating wall thickness. The exhaustive search algorithm seeks for the shortest edge among all edges interconnecting the pixels of the contours of two crypts. The heuristic algorithm computes the distance between the two pixels at the intersection between the contours of the two neighboring crypts and the line connecting their centroids.

Similarly to maximal Feret diameter, we see in Supplementary Fig. 3 that there is a variation on the estimates done using both methods. Here, exhaustive search algorithm provides smaller estimates, and the heuristic, larger for either R or T images. The Hellinger distance between the histograms of Supplementary Figs. 3A and 3B is 0.108 and 0.093, respectively, which shows the great similarity between the distributions in each histogram. The mean value ± standard deviation for the heuristic method is 40.13 μm ± 16.73 μm and 28.28 μm ± 14.92 μm for R and T images, respectively, while for the exhaustive search they are 38.26 μm ± 16.65 μm and 26.06 μm ± 15.64 μm. The relative error, is 0.05 and 0.08 for R and T images, respectively. The estimation of wall thickness using the exhaustive search takes more than 1702 seconds while the use of the heuristic function requires about 3 seconds. The larger time to run the exhaustive search algorithm indicates that the heuristic approach can be very useful when we employ Motiro for the analysis of a large amount of data. Indeed, inspection of the relative error, the Hellinger distance and the mean values allows one to choose the heuristic approach without loss of significant amounts of information. The tissue density is the ratio between the background contour area, denoted by $B$, and the sum of areas of all elliptical contours

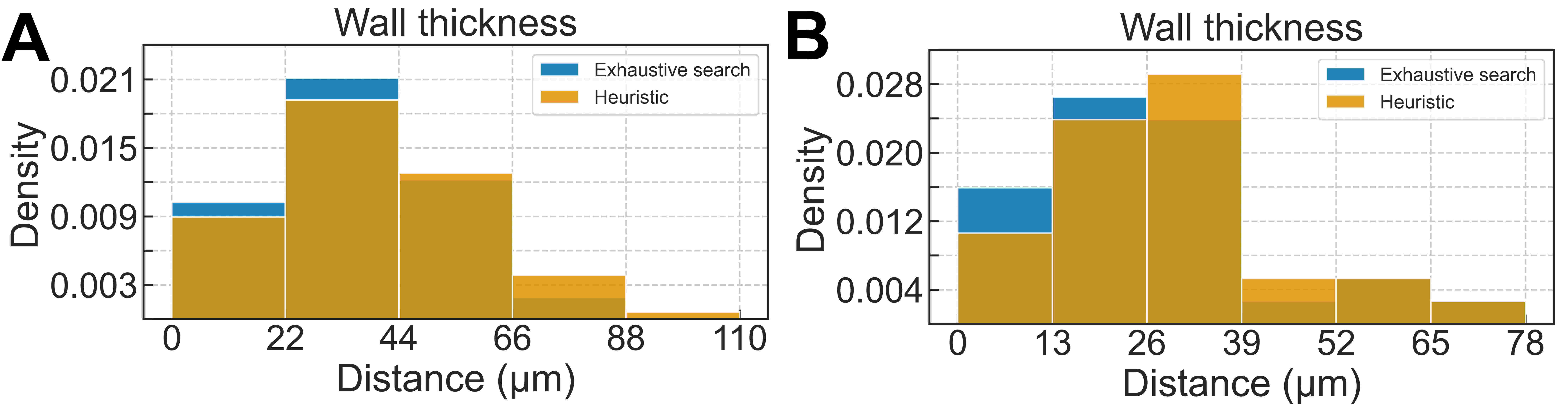

**Supplementary Figure 3: Wall thickness estimation.** Estimated value with exhaustive search and heuristic algorithm for R **(A)** and T **(B)** images

3. **Statistical analysis**

The histograms summarizing the statistics of the morphometric parameters was built using Numpy and Seaborn Python packages. Additionally, we computed the differential Shannon entropy of each histograms as a metrics for the degree of disorder of each parameter of a sample R or T. The larger the differential Shannon entropy is, the closer to a uniform distribution its histogram is, and, consequently, the higher is the disorder of the sample. A Random Variable governed by a uniform distribution has no drift providing an organizational bias that induce its values to be concentrated within a given range (or ranges) while the dynamics of the stochastic process governing the system happens. Such a bias is responsible, for example, for guiding the crypts formation with size and shapes lying within specific ranges during a given phase of the development of the mucosa. For a random variable $X$ assuming real values denoted by $x$, the probability density at $X=x$ is denoted by$p\left( x \right)$. The differential Shannon entropy is defined as $S=-\int p\left( x \right){log}_{2}\left( p\left( x \right) \right)dx$. We estimate the probability density based on the relative frequencies as obtained from our images. Let us consider $N$ bins, with the $i$-th bin limited by $x_{i-1}$ and $x_{i}$. Then we estimate the probability density within the bin as a constant $p\left( x_{i-1}\leq X<x_{i} \right)\approx\frac{f_{i}}{L_{i}}$, where $f_{i}$ and $L_{i}$ are, respectively, the relative frequency and the width of the $i$-th bin. The integral can be written as $S=-\sum\int\frac{f_{i}}{L_{i}}{log}_{2}\left( \frac{f_{i}}{L_{i}} \right)dx$ , and since $f_{i}$ and $L_{i}$ are constants, we write the differential Shannon entropy of a histogram having $N$ bins as:

$S=-\sum f_{i}{log}_{2}\left( \frac{f_{i}}{L_{i}} \right)$.

Additionally, to have a better interpretation of the entropy of the observed values, we set the differential Shannon entropy for a uniform distribution of a continuous random variable defined on an interval of width $L$ which is $S_{0}={log}_{2}\left( L \right)$. Then we define the quantity $\frac{2^{S}}{L}$ to evaluate the disorder of a distribution or histogram relative to its uniform counterpart: the distribution or histogram reflects a higher degree of disorder for $\frac{2^{S}}{L}$ and better organization otherwise. Now, we can estimate the differential Shannon entropy of a histogram for each morphometric parameter to set a measure of the degree of disorder of the image based on their histograms. That will help us on the development of a set of quantitative criteria to classify a colorectal cancer in its early stages.

Additionally, we can use the statistical distance to measure how similar two distributions are. Again, we estimate the probabilities densities based on relative frequencies. There are several metrics for the similarity between two distributions and, in Motiro, we implemented: the Kullback-Leibler divergence, the Jeffrey divergence, the Bhattacharyya distance, the Jensen-Shannon distance and the Hellinger distance(6–8). The latter has been the best one to distinguish among overlaid histograms that we obtained and its formula is given by:

$\text{H(P, Q)=}\sqrt{1-BC\left( p_{i},q_{i} \right)}$

where $P$ and $Q$ are denoting the histograms for a specific morphometric parameter of the image obtained from two sets of images, namely, R and T, and $BC\left( p,q \right)$ is the Bhattacharyya coefficient, defined as:

$BC\left( p,q \right)=\int\sqrt{p\left( x \right)q\left( x \right)}dx$.

The Bhattacharyya coefficient is used for quantifying the overlay between the probability densities governing a continuous random variable. The relative frequencies of the $i$-th range of two histograms having coincident bins are used for estimating $p_{i}$ and $q_{i}$ as we did for estimating the differential Shannon entropy.

4. **Histograms and Statistical analysis with complete data**

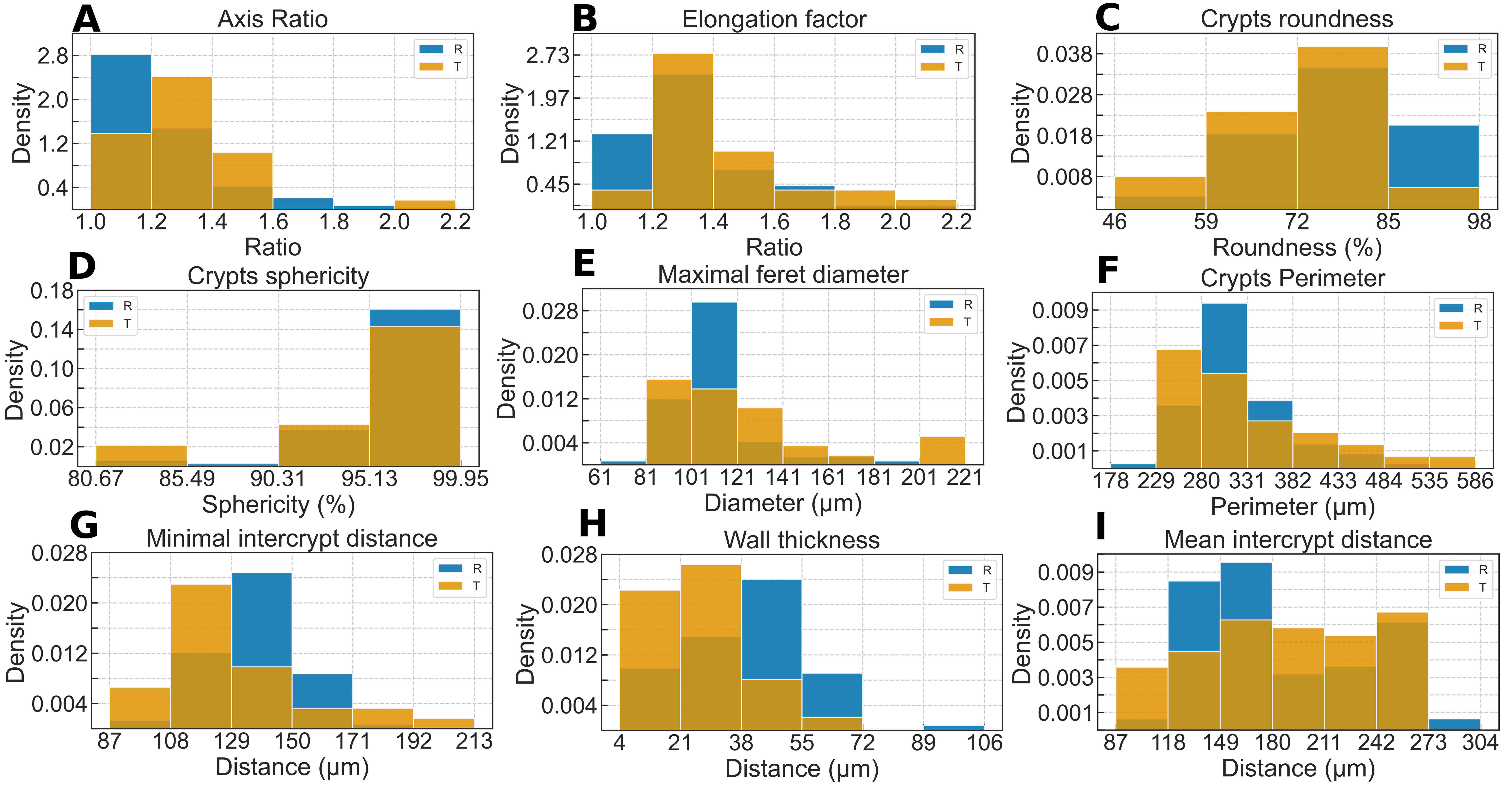
**Supplementary Figure 4:** **Morphometry parameters evaluation.** The blue and beige histograms represent the complete data statistics of the mucosa's morphometric parameters in R and T images, respectively.

***Supplementary Table 2:*** *Complete data statistical analysis of R and T images*

5. **Quantitative comparison between Motiro and Quénéhervé approach**

We compare the results obtained using Motiro with those of Quénéhervé approach(9) (or QA). In QA, Icy Software(10) and ImageJ(11) were used separately, the former for estimating the crypts' axis ratio, elongation factor, Feret diameter, perimeter, roundness, and sphericity, and the latter for estimating the wall thickness, the minimal, and the mean intercrypt distances. Motiro enables a unified analysis of those morphometric parameters and the characterization their statistics in normal tissues. While Motiro enables a non-supervised estimation of the crypts borders (shown in green in article Fig. 2F), QA uses the active cells plugin(12) for semi-automated design of the contours of the crypts as shown by the red circles in article Fig. 2F. On the same set of mosaics, our framework identified 100 crypts while 106 were manually labeled using Icy. Note that the green (red) contours are typically internal (external) to the border of the crypts in article Fig. 2F.

***Supplementary Table 3:*** *A comparison of mean values and standard deviations of the morphometric parameters as estimated using Motiro and Icy Software.*

In the Supplementary *Table 3* we present the mean values (and standard deviation within parenthesis) of the morphometric parameters as estimated by both tools for either R and T stages. The numbers within brackets are the relative error on the estimation of the means, as given by $\left| \mu_{F}-\mu_{I} \right|/{Max}\left( \mu_{F},\mu_{I} \right)$, where $\mu_{F}$ and $\mu_{I}$ are the means obtained using Motiro and QA, respectively. $Max\left( . \right)$ returns the greatest of its arguments. The estimation of the dimensionless (relative) parameters axis ratio and elongation factor are the most similar ones, while sphericity has the largest relative errors, on both R and T stages. That result is consistent with the bias of both methods for estimating the crypts' contour and helps to quantify the uncertainty on the analysis when using either method.

The estimation of the wall thickness, minimal and mean intercrypt distances by the ImageJ Nearest Distances (ND) Plugin(13) demands user interaction. It receives a user produced binary image as input and estimate the contour of the crypts as ellipses. Then the algorithm estimates a regular graph in which the centroid of the crypts are the vertices and the edges connecting two centroids of neighboring crypts set the interconnection of the graph. The user sets the degree of the graph (number of closest neighbors of each crypt) and the lengths of the edges are used for estimating the intercrypt parameters. We set the degree of the crypt's graph as four, as that was the average value estimated using our algorithm.

***Supplementary Table 4:*** *Mean and standard deviation comparison between Motiro and ImageJ Plugin.*

*Supplementary Table 4* shows the mean values (and standard deviations) of the intercrypt parameters estimated with Motiro and QA using videos of the R and T stages. While minimal intercrypt distance is approximately the same for estimates using both methods, Motiro estimates larger values for mean intercrypt distance than QA. That is because Motiro determines the graph's neighborhood by determining whether the crypt's centroids lie within a fixed radius. The fixed length can be set accordingly with physical properties of the intercrypt's interaction such as β-Catenin diffusion within the mucosa(14). In that case, it is fair to expect that the degree of the graph will not be fixed because of the small variations on the crypt's distribution within the mucosa. Notice, however, that the possibility of setting more than four neighbors induces a bias since the fifth crypt is always more distant than the fourth one.

In the T images, the degree of the graphs has mode equals to four and the mean equals to five, and the crypts are more spread than in the R images whose mode and mean of the degrees of the crypt's graphs degrees are both equal to four. Since we just evaluated two videos, we interpret the difference on the degrees of the graphs in the T and R images as just a bias. The largest relative error among the estimates of the intercrypt parameters occurs for wall thickness because the two approaches differ on their calculation methods. QA computes the distance between crypts one and two with centroids at $\left( X_{1},Y_{1} \right)$ and $\left( X_{2},Y_{2} \right)$, respectively, using the formula $d=\sqrt{\left( Y_{2}-Y_{1} \right)^{2}+\left( X_{2}-X_{1} \right)^{2}}-\left( r_{1}+r_{2} \right)$, where $r_{1}+r_{2}=\frac{a_{1}+a_{2}+b_{1}+b_{2}}{2}$ is given in terms of the semi-major and semi-minor axis of the ellipses. This approach will systematically overestimate the wall thickness. Such a bias is prevented in Motiro as the wall thickness is estimated as the minimal distance between the contour of two neighboring ellipses. Note that the input binary mosaic for application of both methods was built using Motiro.

6. **Runtime benchmark between Motiro and Quénéhervé approach**

The machine used to compare the computing time of the two methods was a laptop with the following specifications: a CPU Intel® Core™ i7-8565U 1.80 GHz Quad-Core, a GPU Nvidia GeForce MX150 2GB GDDR5, a RAM memory of 32GB DDR4 with the Linux Operation System Ubuntu 18.04.5 LTS(x64). The Motiro packages versions are Python(3.6.9), Seaborn(0.10.1), Numpy(1.19), and OpenCV(4.3.0). QA used software versions are ImageJ(1.51q) with Java 1.8 and Icy(2.0.3.0). The time measure of Motiro was automatically evaluated by *timeit* Python package and the spend time in QA was measured with a digital chronometer whereby the software were fully load and the images were opened before the start the digital chronometer to evaluate the time elapsed to analysis and data storage.

Here, we compare the runtime for Motiro and QA to estimate the morphometric parameters. The estimates of the morphometric parameters can be divided into three stages. *1)* The estimate of the elliptic contours of the crypts; *2)* The estimate of the crypt's morphometric parameters; *3)* The estimate of the intercrypt distances.

In QA's approach a mosaic produced using CellVizio software is used as input. The stage *1)* is carried out using Icy to open mosaic image, annotating the crypts and estimating their contours as ellipses. Then, Active Cells Icy plugin is used for fine tuning the estimates of the crypts' contours. The stage *2)* is done applying the built-in morphometry Icy functions. The stage *3)* is done by opening the image obtained at stage *1)* using ImageJ and ND plugin to determine the regular graphs formed by the crypts. Note that the degree of the regular graphs is set by the user.

In Motiro the input is the video produced using pCLE to our script written using Python. In stage *1)* our script uses Python's library OpenCV for dismantling the video into frames and calls ImageJ's plugin Register Virtual Stack Slices for mosaic construction. Then OpenCV is used for pre-processing, segmentation, and estimating the elliptic contours of the crypts. The stage *2)* is done applying OpenCV and NumPy functions for estimating the crypts' morphometric parameters. The stage *3)* also uses function from OpenCV and NumPy for estimating the intercrypts' parameters.

For stage *1)*: Motiro estimates 100 crypts in R and T images in 155 seconds, where we neglect the time spent on the construction of the mosaic; QA estimates 106 in R and T images in 874 seconds.

For stage *2)*: Motiro estimates the morphometric parameters of the crypts in 3 seconds, if we use the heuristic estimate for the maximal Feret diameter, or in 205 seconds, if the exhaustive search method is employed; QA estimates those parameters as the elliptic contours are estimated.

For stage *3)*: the image with estimates of the elliptic contours done by Motiro at stage *1)* is used as input for both methods. Motiro uses Numpy and OpenCV for estimating the intercrypt parameters in 5 seconds (or 1703 seconds) if the heuristic (or the exhaustive search) algorithm is used. QA uses ND plugin and estimates of the intercrypt parameters take 54 seconds.

The total time for completing application of Motiro using the heuristic algorithms was 163 seconds or 2063 seconds using exhaustive search, and we emphasize that Motiro does its analysis using a single script in a non-supervised fashion. The total time for completing application of QA is 928 seconds in a partially supervised fashion. The analysis using QA can be faster than using Motiro if we employ the exhaustive search method. However, as we have shown before, the heuristic estimates are in sufficiently good agreement with the ones done using exhaustive search. Therefore, it is fair to assume that Motiro is 5.7 times faster than QA when applied for the same mosaics used here. Besides, QA requires the use of more than one software and user interaction. Hence, it is fair to conclude that our approach represents a great improvement when compared with the previous analysis.

**SUPPLEMENTARY REFERENCES**

1.	Applied Mathematics and Biological Physics Laboratory: Endomicroscopy analysis [Internet]. [cited 2020 Jul 8] Available from: https://github.com/amphybio/endomicroscopy-analysis

2.	Bradski G, Kaehler A. OpenCV. Dr Dobb’s J. Softw. Tools 2000;3

3.	Arganda-Carreras I, Sorzano COS, Marabini R, *et al.* Consistent and Elastic Registration of Histological Sections Using Vector-Spline Regularization. In: Beichel RR, Sonka M, editor(s). Computer Vision Approaches to Medical Image Analysis. Berlin, Heidelberg: Springer Berlin Heidelberg; 2006. p. 85–95.

4.	Lowe DG. Object recognition from local scale-invariant features [Internet]. In: Proceedings of the Seventh IEEE International Conference on Computer Vision. Kerkyra, Greece: IEEE; 1999. p. 1150–1157 vol.2.[cited 2020 Jul 17] Available from: http://ieeexplore.ieee.org/document/790410/

5.	Jain AK, Murty MN, Flynn PJ. Data clustering: a review. ACM Comput. Surv. 1999;31:264–323.

6.	Chung JK, Kannappan PL, Ng CT, *et al.* Measures of distance between probability distributions. J. Math. Anal. Appl. 1989;138:280–292.

7.	Gibbs AL, Su FE. On Choosing and Bounding Probability Metrics. Int. Stat. Rev. 2002;70:419–435.

8.	Deza MM, Deza E. Encyclopedia of Distances. Berlin, Heidelberg: Springer Berlin Heidelberg; 2009.

9.	Quénéhervé L, David G, Bourreille A, *et al.* Quantitative assessment of mucosal architecture using computer-based analysis of confocal laser endomicroscopy in inflammatory bowel diseases. Gastrointest. Endosc. 2019;89:626–636.

10. Chaumont F de, Dallongeville S, Chenouard N, *et al.* Icy: an open bioimage informatics platform for extended reproducible research. Nat. Methods 2012;9:690–696.

11.	Schneider CA, Rasband WS, Eliceiri KW. NIH Image to ImageJ: 25 years of image analysis. Nat. Methods 2012;9:671–675.

12. Delgado-Gonzalo R, Uhlmann V, Schmitter D, *et al.* Snakes on a Plane: A perfect snap for bioimage analysis. IEEE Signal Process. Mag. 2015;32:41–48.

13. Haeri M, Haeri M. ImageJ Plugin for Analysis of Porous Scaffolds used in Tissue Engineering [Internet]. J. Open Res. Softw. 2015;3[cited 2020 Aug 26] Available from: http://openresearchsoftware.metajnl.com/articles/10.5334/jors.bn/

14. Murray PJ, Kang J-W, Mirams GR, *et al.* Modelling Spatially Regulated β-Catenin Dynamics and Invasion in Intestinal Crypts. Biophys. J. 2010;99:716–725.

***Supplementary Table 2:*** *Complete data statistical analysis of R and T images*

|  |  | **Axis Ratio** | **Elongation Factor** | **Roundness(%)** | **Sphericity(%)** | **Maximal Feret diameter(μm)** | **Perimeter(μm)** | **Minimal intercrypt distance(μm)** | **Wall thickness(μm)** | **Mean intercrypt distance(μm)** |
| --- | --- | --- | --- | --- | --- | --- | --- | --- | --- | --- |
| **R** | **mean (*±std*)** | 1.21 (±0.18) | 1.33 (±0.21) | 77.0 (±10.4) | 96.6 (±3.5) | 112.0 (±19.4) | 318.6 (±53.9) | 137.9 (±15.1) | 40.1 (±16.7) | 185.2 (±47.5) |
|  | **Degree of disorder** | 0.494 | 0.618 | 0.820 | 0.488 | 0.408 | 0.519 | 0.531 | 0.651 | 0.748 |
| **T** | **mean (*±std*)** | 1.32 (±0.21) | 1.44 (±0.24) | 71.3 (±10.1) | 94.9 (±4.5) | 121.4 (±35.6) | 328.4 (±83.2) | 127.8 (±27.8) | 28.3 (±14.9) | 189.6 (±52.1) |
|  | **Degree of disorder** | 0.526 | 0.626 | *0.769* | 0.566 | 0.607 | 0.649 | 0.700 | *0.509* | 0.840 |
|  | **Statistical Distance** | 0.309 | 0.223 | 0.206 | 0.143 | 0.341 | 0.243 | 0.340 | 0.315 | 0.230 |

***Supplementary Table 3:*** *A comparison of mean values and standard deviations of the morphometric parameters as estimated using Motiro and Icy Software.*

|  |  | **Axis Ratio** | **Elongation factor** | **Roundness(%)** | **Sphericity(%)** | **Maximal Feret diameter(μm)** | **Perimeter(μm)** |
| --- | --- | --- | --- | --- | --- | --- | --- |
| **R** | **Motiro**  **mean (*±std*)** | *1.21 (±0.18)* | *1.33 (±0.21)* | *77.0 (±10.4)* | *96.6 (±3.5)* | *112.0 (±19.4)* | *318.6 (±53.9)* |
|  | **Icy**  **mean (*±std*)** | *1.21 (±0.14)* | *1.27 (±0.14)* | *61.7 (±4.3)* | *64.4 (±0.6)* | *149.7 (±13.9)* | *389.4 (±32.7)* |
|  | **Relative**  **Error** | 0.00 | 0.045 | 0.198 | 0.335 | 0.251 | 0.181 |
| **T** | **Motiro**  **mean (*±std*)** | *1.32 (±0.21)* | *1.44 (±0.24)* | *71.3 (±10.1)* | *94.9 (±4.5)* | *121.4 (±35.6)* | *328.4 (±83.2)* |
|  | **Icy**  **mean (*±std*)** | *1.38 (±0.24)* | *1.46 (±0.24)* | *53.1 (±8)* | *62.9 (±1.7)* | *132.7 (±12.6)* | *340.0 (±33.7)* |
|  | **Relative**  **Error** | 0.004 | 0.013 | 0.255 | 0.337 | 0.085 | 0.034 |

***Supplementary Table 4:*** *Mean and standard deviation comparison between Motiro and ImageJ Plugin.*

|  |  | **Minimal intercrypt distance(****μm)** | **Mean intercrypt distance(μm)** | **Wall thickness(μm)** |
| --- | --- | --- | --- | --- |
| **R** | **Motiro**  **mean (*±std*)** | 137.9 (±15.1) | 185.2 (±47.5) | 40.1 (±16.7) |
|  | **ImageJ**  **mean (*±std*)** | 137.8 (±15.1) | 184.7 (±23.9) | 89.3 (±29.5) |
|  | **Relative**  **Error** | 0.00 | 0.002 | 0.55 |
| **T** | **Motiro**  **mean (*±std*)** | 127.8 (±18.7) | 189.6 (±52.1) | 28.3 (±14.9) |
|  | **ImageJ**  **mean (*±std*)** | 120.6 (±18.8) | 177.6 (±25.3) | 70.6 (±20.5) |
|  | **Relative**  **Error** | 0.056 | 0.063 | 0.599 |
